# Prevalence of TBI in an SUD Population

**DOI:** 10.1101/2025.07.25.25332207

**Authors:** Ryan Coen, Alyssa Lester, Zaccardi Muniz, Lura Leech, Amanda Acord-Vira, SueAnn Woods, Steven Wheeler

**Affiliations:** Division of Occupational Therapy, West Virginia University OTH 697: Research Capstone

**Keywords:** Traumatic Brain Injury, Substance Use Disorder, routines, support system, functional independence, risk factors, cognition, impulsivity, mood regulation, relationship

## Abstract

**Methodology:** *Study Design:* This study utilized a cross-sectional design that assessed patients with a history of TBI (TBI) admitted to a substance use disorder (SUD) treatment program at a Recovery Point in West Virginia. This design provided a point-in-time measure of TBI prevalence in this population and allowed for finding associations between TBI and substance use.

*Participants:* The participant sample that was selected included individuals who were 18 years or older and confirmed that they had been admitted to the substance use disease treatment program. Those eligible individuals were recruited through admissions; the inclusion criteria included participants to be screened for a potential history of TBI using the Ohio State University TBI identification method (OSU-TBI-ID) (Bogner et al., 2009). The only participants included in this study were individuals who screened positive for TBI and were included in the study cohort demographic information, which included age, gender, and substance use history. This information was collected as part of the mental health assessment administered by trained non-physician personnel.

*Data Collection:* Data were collected during the standard intake procedure at Recovery Point. The ASAM provided essential contextual data for each participant, which included mental health history and substance use patterns.

*Data Analysis:* The de-identified data analysis obtained from the ASAM was conducted at the University of Arkansas. The Center for Excellence in Disabilities facility allowed the research team to complete their data entry, and the pooled data were analyzed using descriptive statistics and SPSS software.

*Ethical Considerations:* The Institutional Review Board (IRB) approval for the study was obtained from West Virginia University, and the study adhered to the ethical principles outlined in the Declaration of Helsinki. Informed consent was obtained from all participants before participating, and all data were kept confidential to protect participants’ privacy. This study included some limitations, which include self-reports for substance use and TBI history. Future research could benefit from longitudinal designs to further explore these relationships.

## Prevalence of TBI and SUD Population

TBI has a great impact on individuals and their family’s lives. There are millions of individuals each year who suffer from a TBI, and that number continues to rise, especially in young adults and older adults. It is one of the most common causes of death, economic burden, and disability in the world (Khellaf et al. 2019). TBI can be categorized into mild, moderate, and severe according to the Glasgow Coma Scale. A mild TBI is the most common type, also known as a concussion. Most reported TBIs consist of mild TBI, which often leads to them being underreported or underestimated, and leads to disparities in their true impact on affected individuals. TBI can cause various disabilities, including physical, cognitive, and psychosocial domains. Some leading causes of TBI include falls, motor vehicle accidents, and sports-related injuries, with each having its own set of risk factors and recovery time. Khellaf et al. (2019) literature review emphasized future implications for head trauma management. The topics that were covered included proposed potential neuro-intensive care targeted protocol therapy, evidence on the role of decompressive craniotomy, and new drug therapies to get plenty of ideas on the table. Howe et al. (2024) explained the prevalence of post-traumatic headaches and the 12-month post-TBI predictors, which also included valuable insights on the post-injury symptoms after TBI.

Haarbauer-Krupa et al. (2021) is one of many articles that have contributed to information about the rates of TBI in recent years. They conducted a comprehensive review of the chronic effects of TBI. They also discovered that TBIs caused by mid-range impacts are difficult to identify but common. Machamer et al. (2022) identified symptoms that were experienced over the first year following a TBI, which included a list of persistent and transient symptoms that were reported post-injury. They also emphasized that the knowledge of the epidemiology of characteristic pics of TBI is crucial in enhancing the quality of life of individuals with TBI.

### TBI Symptoms

TBI effects vary in each person’s physical, cognitive, emotional, and behavioral state. Symptoms include, but are not limited to, headaches, dizziness, nausea, and fatigue. The physical symptoms are caused by the injury directly impacting the brain, which requires immediate medical attention and intervention (Haarbauer-Krupa et al., 2021). A moderate to severe TBI can cause long-term adverse effects after five years; these effects include moderate to severe disability, loss of employment, increased use of illicit drugs, or misuse of alcohol (Centers for Disease Control and Prevention [CDC], 2024).

#### Cognitive Symptoms

Deficits from a TBI can include shortened attention span, memory, executive functioning, and information processing speed (Khellaf et al., 2019). Individuals’ day-to-day activities can also be impacted by problems, such as concentrating, organizing, and solving problems. TBI also impacts communication skills, which results in challenges with receptive and expressive language abilities that impact socialization and pragmatic communication. Physical symptoms are usually the first to appear after the injury, and the cognitive symptoms are least likely to vary over time or diminish over time based on the severity (Machamer et al., 2022).

Wilson et al. (2021) discovered that those individuals who self-reported a full cognitive recovery after a history of TBI had utilized compensatory strategies to remain independent and not return to their baseline cognitive functioning. For example, this means the individual displayed a slow processing speed, which meant completing a task at a slower rate. Compensatory strategies cause a degenerative disease in later age due to being overused in overstressed environments. TBI affects each person differently, which means a wide range of physical, cognitive, emotional, and behavioral difficulties.

#### Emotional and Behavioral Symptoms

TBI can dramatically change a person’s emotional and behavioral state permanently. There are signs and symptoms of these changes, which include depression, anxiety, irritability, and emotional lability. These, in turn, impact the individual’s quality of life and social relationships (Howe et al., 2024). Individuals with a known history of TBI experience higher levels of cognitive distress regarding social determinants of health (SDoH), which are commonly correlated to a lower quality of life when adversely impacted (Chan et al., 2022). Ineson et al. (2023) established that TBI causes behavioral sequelae, including impulsivity, disinhibition, and emotional dysregulation, which increases the risk of suicidal ideation in violent crimes. These symptoms can cause major strains in the individual’s work and relationships.

TBIs affect not only the person directly but also the person’s family, caregivers, and other support systems (Khellaf et al.,2019). TBIs are unique to each person, and they require a more holistic approach to both support and rehabilitation. The approach requires an interdisciplinary and collaborative effort in the treatment to address the full range of needs for the TBI population in hopes of improving the recovery and continuation of life in the community. Awareness of the diversity of TBIs allows healthcare professionals to address the evolving needs of TBI survivors while promoting their well-being and quality of life.

### Prevalence of SUD

SUD cases have increased tremendously and accounted for 147.5 million cases in 2015 (Prom-Wormley et al., 2017). This disorder affects all the individual’s activities of daily living (ADL) in almost every aspect of their life (Alkhawaldeh et al., 2023). Some comorbidities are often associated with substance use disorder, which includes anxiety, depression, and TBI. TBI and SUD diagnosis often co-occur because SUD alters the individual’s way of life and increases the performance of high-risk behaviors (Merkel et al., 2017). Awan et al. (2020) discovered that contextual factors play a role in determining which individuals are at higher risk for substance use disorder. For instance, people who use substances at a younger age are more susceptible to developing this disorder. These risk factors increase the potential of knowing when this disorder may develop and show which demographics are more susceptible to the legal use of legal and illegal substances at what age.

Genetics may also play a part in whether a person will develop substance use disorder, in which children with parents who were diagnosed with SUD are at greater risk (Prom-Wormley et al., 2017). The genes that enable addictiveness can cause this disorder, which impacts the youth and creates a vicious cycle of generational substance use disorder. Due to the reduced executive functioning in increased risk-taking behavior that occurs for both TBI and SUD, there is a greater prevalence of individuals experiencing these diagnoses simultaneously.

### Characteristics of SUD

SUD signs and symptoms can vary by person, but they can also be described in a way that aids in diagnosis and classification. For example, prescription opioids are associated with psychological symptoms that include depression, anxiety, and post-traumatic stress disorder (PTSD) (Starosta et al., 2021). SUD can be described as an individual who uses any illegal substance, uses prescribed medication for longer than prescribed, and admits that the substance takes up much of their time (Rhemtulla et al., 2016). The use of substances can also result in withdrawal effects, which influence an individual’s continuing consumption of the substance. SUD can be categorized into severity ranks, such as mild, moderate, and severe. Mild occurs when two to three of the criteria are met, moderate 1/4 to five symptoms are met, and severe when six of the criteria are met (Hasin et al., 2013).

### Relationship between TBI and SUD

Starosta et al. (2021) noted that individuals who have obtained a TBI are at increased risk for developing an out-of-control use of illicit substances. Individuals with a history of SUD are at increased vulnerability to a TBI, and so this association highlights the potential relationship between TBI and SUD. It also demonstrates how the two disorders can affect each other, which leads to worsened symptoms and outcomes. An individual who sustained a TBI is possibly more likely to begin using or selling drugs as a way of coping with the physical, cognitive, and emotional symptoms of their injury. There is a risk of developing SUD, which is heightened due to impacted executive functioning resulting from TBI. Conversely, individuals with SUD increase their chance of having TBIs due to behaviors such as poor decision-making, risky behaviors, and getting into accidents or fights.

Grossbard et al. (2017) focused on veterans with TBIs and the high prevalence of alcohol misuse and emphasized the need for early identification and interventions that target alcohol misuse. There is an association between prescription opioid medications and psychological symptoms among individuals with TBI, in turn increasing substance use disorders within this population (Starosta et al. 2021). Awan et al. (2020) explained how people with a TBI history are more susceptible to injuries related to substance abuse and have poor outcomes following their injury.

Substance use may exacerbate TBI symptoms and impact the management of the symptoms, which may lead to non-compliance with treatment and delay in the individual’s functional independence. Repeated TBIs have been associated with comorbid psychiatric symptoms and SUD, especially in more severe cases (McHugo et al., 2017). Starosta et al. (2021) argued that SUD can impact the diagnosis of symptoms that are secondary to TBI, which in turn creates an underestimated effect of the injury, hindering the proper treatment for the individual.

Healthcare providers should be cautiously aware of the link between SUDs and TBIs to create tailored treatment approaches, treat cognitive dysfunction, and incentivize healthy coping mechanisms to help the individual achieve functional outcomes and improve overall wellness (Starosta et al., 2021).

### Implications of OT in TBI and SUD

Occupational therapy is found to be very effective when it comes to helping individuals with TBI and or SUD back to participating in their daily routines that have been disrupted because of their conditions (Alkhawaldeh et al., 2023; Wheeler et al., 2016). TBI and SUD have various severities that disrupt routines and occupational performance, and therefore, occupational therapy can be useful in remediating or compensating for these diagnoses. Individuals with TBI tend to spend most of their time in acute care settings to promote medical stability (Alkhawaldeh et al., 2023). This article also highlights the importance of early OT intervention in reducing the length of stay in acute care of patients who have suffered moderate to severe TBI.

Having increased knowledge in this area enables the OT to advocate for their clients, assess them, and take the most determined actions in their care (Alkhawaldeh et al., 2023). An occupational therapist is equipped with skills that allow them to relate to their clients’ concerns and, therefore, enable them to learn how this disorder impacts their occupational roles. Even though occupational therapy is an effective treatment option for both SUD and TBI, OTs frequently feel ill-equipped to do so (Matilla et al., 2022). The qualitative research discovered that of 116 practitioner participants, 84 of them had no formal training with individuals facing substance use disorder, showing that most of the practitioners are unprepared to work with individuals of this population (Alkhawaldeh et al., 2023).

While many OT practitioners may feel less prepared to treat individuals with SUD, it is found that occupational-based interventions can be very effective for these individuals’ recovery (Matilla et al., 2022; Ryan et al., 2023). Ryan et al. (2023) conducted a cohort study that aimed to investigate the effectiveness of enhancing occupational balance. This was achieved through volition and habituation OT sessions that promote mental health in individuals with SUD. It is up to the healthcare team to design a personalized treatment plan that addresses both the short-term and long-term goals of this population (Rosenthal et al., 2023).

### Purpose of the Study

The purpose of this study is to examine the prevalence and association between TBI and substance use disorder. This association is complex and challenging due to the overlap of confounding variables between these conditions. It is quite challenging to determine whether SUD is more likely to be the cause of TBI or whether TBI is more likely to be the precursor of substance use disorder. SUD can be characterized by risk-taking, which in turn results in a greater chance of sustaining a TBI, while individuals with TBI may use substances to cope with the symptoms associated with their injury. Therefore, highlights the intricate relationship and the importance of this study in exploring the complex relationship between TBI and SUD diagnosis.

## Methods

The data was collected from Recovery Point, which is a non-profit organization, and the employees utilized the Ohio State University TBI Identification Method (OSU-TBI-ID) to screen for a possible history of TBI (Bogner et al., 2009). This screener consists of yes or no questions that indicate whether the individual has had any form of head injury or concussion. The individuals who received a positive screen were included in the study’s participants. The Mental Health Assessment by a non-physician (ASAM) is used as part of the normal program admission intake process. The data from the ASAM was de-identified by an impartial third party and was analyzed by the research team using SPSS. This data was then further analyzed and reported as pooled data via descriptive statistics.

### Participants

The participants were recruited from four different recovery point centers that address substance use disorder in West Virginia. The inclusion criteria for the participants include being English-speaking, ages 18 and older, a positive screen for the OSU-TBI-ID, and the use of one or more substances. The exclusion criteria for the participants include negative screens for the OSU-TBI-ID, inability to complete the screener, and ASAM.

### Measures

The measures of the study were collected through the Ohio State University Traumatic Brain Injury Identification Method (OSU TBI-ID) and the Mental Health Assessment by a Non-Physician (ASAM). Both measures are standardized and were given by the staff at Recovery Point. The OSU TBI-ID is a screening tool that takes approximately 2-5 minutes to complete and identifies individuals who have had a diagnosed brain injury and individuals who could potentially have an undiagnosed brain injury. The questions were designed to flag the individuals who have had head trauma in the past. The responses of this screener can be obtained through the participant or via proxy.

The ASAM is a questionnaire that addresses a multitude of different contextual and personal factors for the participant. This includes personal history, mental health, substance use and frequency, triggers, and social history. This assessment allowed for semi-structured elaborations on all these factors. These responses allowed for a better understanding of the participants with areas of concern and their own personal history. For this study, the data collected from this assessment allowed for a more comprehensive understanding of how the participants’ TBI and SUD may impact other aspects of life.

### Data Analysis

The data was collected from Recovery Point and sent to the West Virginia Center of Excellence and Disability via a secure file and was deidentified by a third-party employee who was not affiliated with this research. Once the data were deidentified, researchers utilized Statistical Package for the Social Sciences (SPSS) to collect ongoing data. Descriptive statistics were run to analyze the current data. This is a currently ongoing project and will continue at the West Virginia Center of Excellence and Disability.

## Results

Table 1 displays demographic data for study participants (n=483). Males were the most frequent participants at 70.4%. White was the most reported race, accounting for 88.2% of participants. Those who identified as heterosexual accounted for 90.9% of participants, with the second most reported sexual orientation being Bisexual (7.7%). The highest reported status for Marital/Relationship status was Single (55.7%), which was reported significantly more than the second highest reported status, Divorced (18.2%). The most common self-reported causes of traumatic brain injury from the TBI-OSU-ID are given in Table 2. Motor Vehicle Accidents (n=184) were the leading cause of TBI, followed by Violence (n=149). These causes of TBI were significantly higher than the third-highest reported cause, Sports (n=41). Difficulty with feelings, hanging around the wrong places, and boredom were regarded as being the largest triggers for alcohol and/or substance use, with the frequency of these data points being over 70% for all three in Table 3. Table 4 provides data on the negative impact of substance use on daily functions. The highest reported negative impact was on relationships, with a frequency of 91.3% of individuals marking “Yes” for that data point. Table 5 displays the frequency of substances used. Tobacco/nicotine, marijuana, alcohol, and methamphetamine had a frequency response rate of over 90%, compared to the fifth most frequently used substance, crack/cocaine, with a frequency of 74% of responses.

**Table 1.**
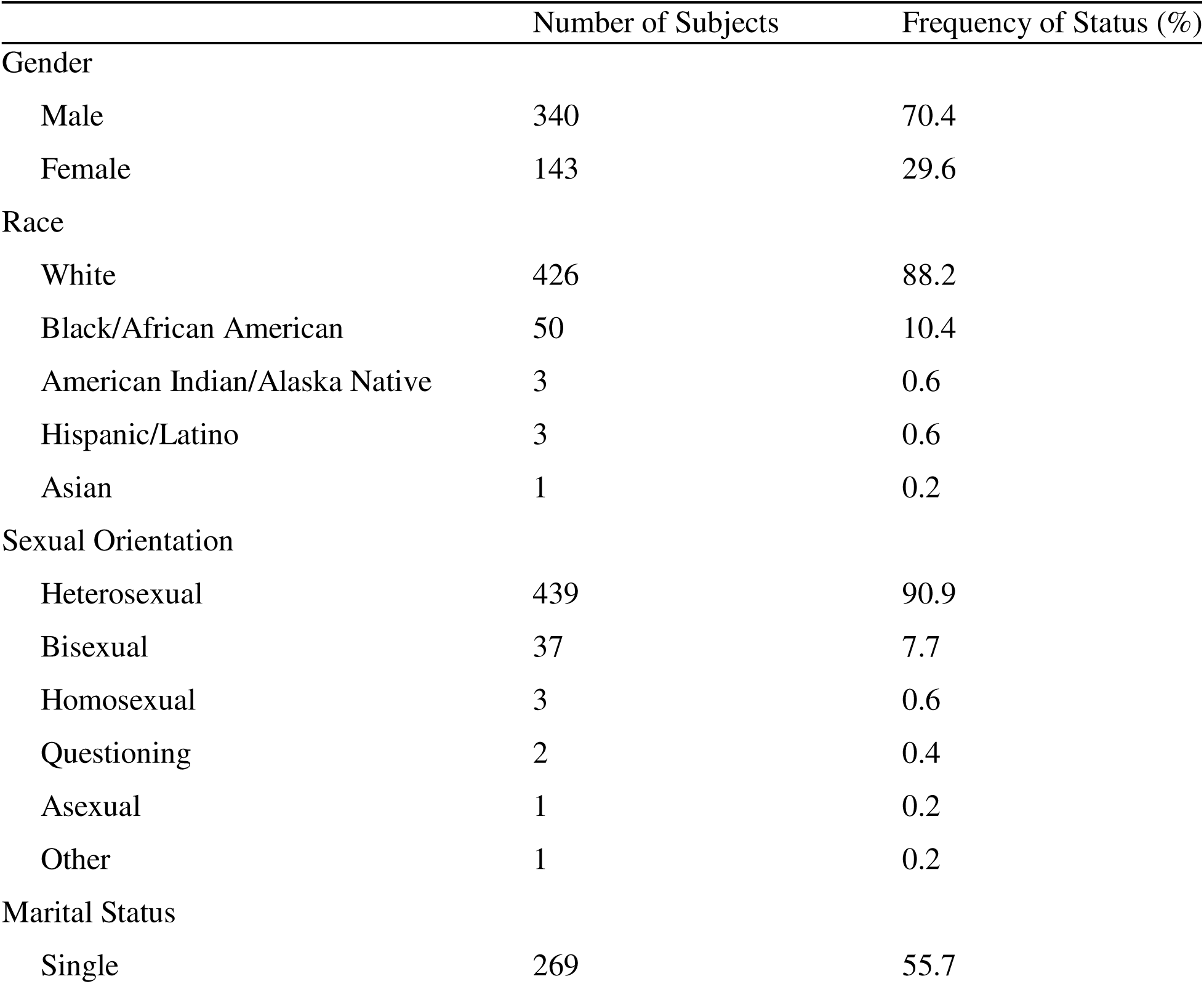
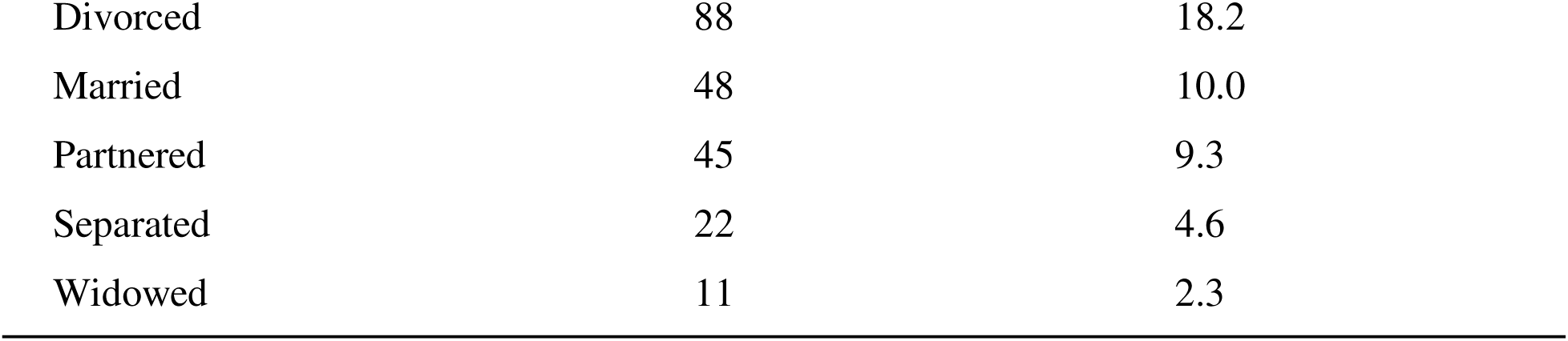
Demographic Characteristics of Participants.

|  | Number of Subjects | Frequency of Status (%) |
| --- | --- | --- |
| Gender |  |  |
| Male | 340 | 70.4 |
| Female | 143 | 29.6 |
| Race |  |  |
| White | 426 | 88.2 |
| Black/African American | 50 | 10.4 |
| American Indian/Alaska Native | 3 | 0.6 |
| Hispanic/Latino | 3 | 0.6 |
| Asian | 1 | 0.2 |
| Sexual Orientation |  |  |
| Heterosexual | 439 | 90.9 |
| Bisexual | 37 | 7.7 |
| Homosexual | 3 | 0.6 |
| Questioning | 2 | 0.4 |
| Asexual | 1 | 0.2 |
| Other | 1 | 0.2 |
| Marital Status |  |  |
| Single | 269 | 55.7 |
| Divorced | 88 | 18.2 |
| Married | 48 | 10.0 |
| Partnered | 45 | 9.3 |
| Separated | 22 | 4.6 |
| Widowed | 11 | 2.3 |

**Table 2.** Cause of Traumatic Brain Injury.

| Cause of TBI | Number of Cases | Frequency of Cause (%) |
| --- | --- | --- |
| Motor Vehicle Accident | 184 | 38.2 |
| Violence | 149 | 30.9 |
| Sports | 41 | 8.5 |
| Falls | 35 | 7.3 |
| Other | 25 | 5.2 |
| Recreational Vehicle | 20 | 4.1 |
| Bicycle | 13 | 2.7 |
| Workplace | 9 | 1.9 |
| Pedestrian | 6 | 1.2 |

**Table 3.**
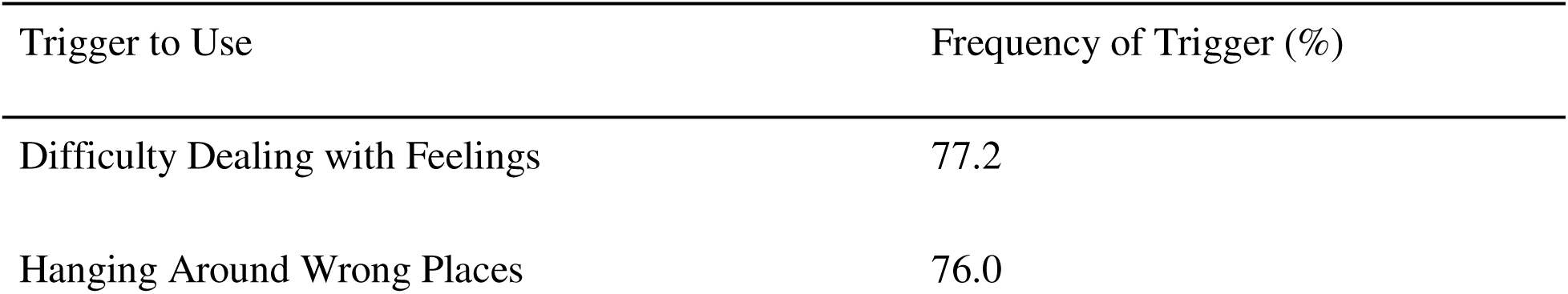
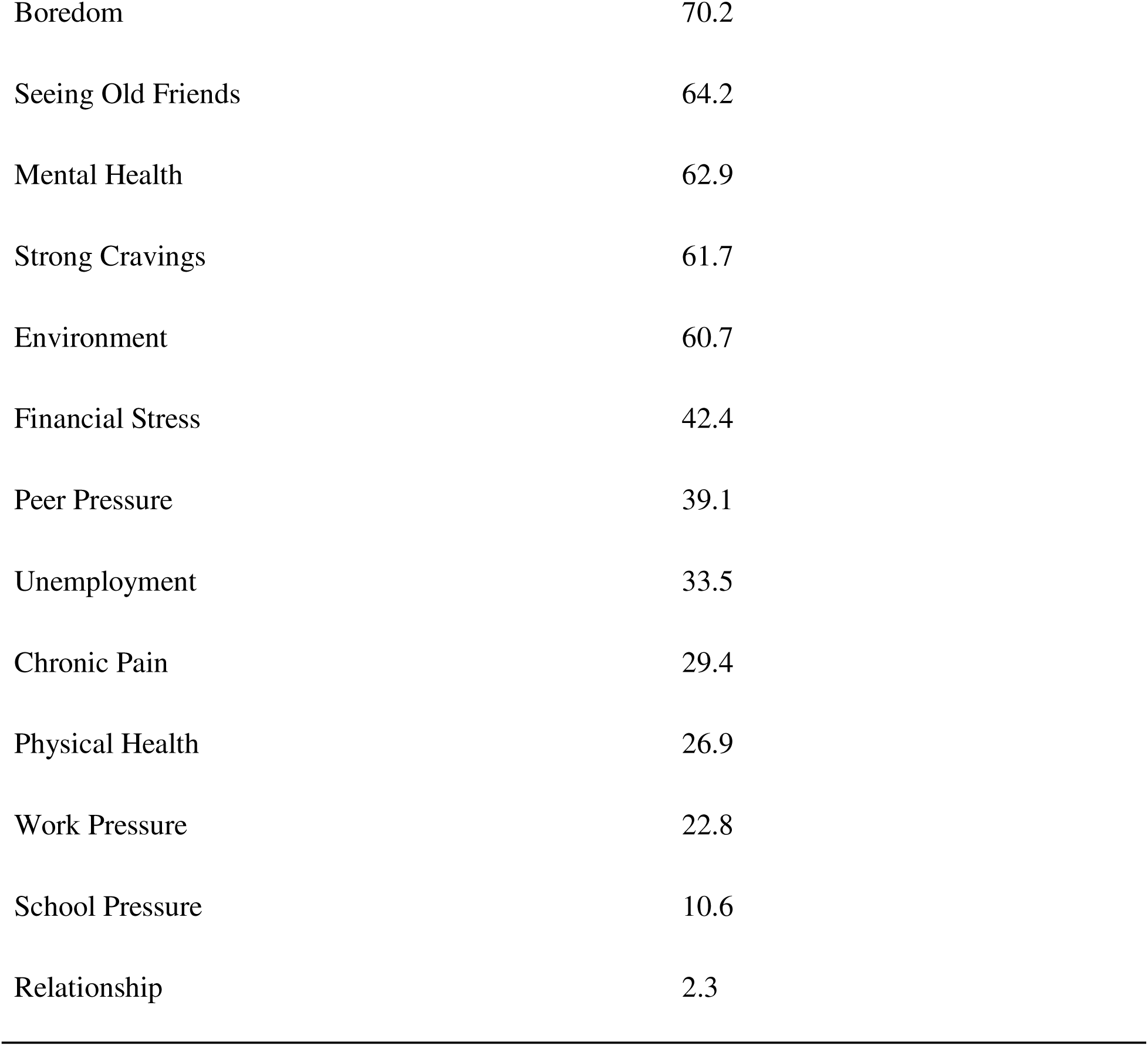
Sel-Reported Triggers to Use Alcohol and/or Substances.

| Trigger to Use | Frequency of Trigger (%) |
| --- | --- |
| Difficulty Dealing with Feelings | 77.2 |
| Hanging Around Wrong Places | 76.0 |
| Boredom | 70.2 |
| Seeing Old Friends | 64.2 |
| Mental Health | 62.9 |
| Strong Cravings | 61.7 |
| Environment | 60.7 |
| Financial Stress | 42.4 |
| Peer Pressure | 39.1 |
| Unemployment | 33.5 |
| Chronic Pain | 29.4 |
| Physical Health | 26.9 |
| Work Pressure | 22.8 |
| School Pressure | 10.6 |
| Relationship | 2.3 |

**Table 4.** Self-Reported Negative Impact of Substance Use on Daily Functions.

| Impact Area | Frequency of Negative Impact (%) |
| --- | --- |
| Relationships | 91.3 |
| Legal Matters | 89.0 |
| Finances | 81.6 |
| Mental Health | 78.9 |
| Work | 77.0 |
| Self-Esteem | 70.6 |
| Physical Health | 69.6 |
| Recreational Activities | 68.7 |
| Everyday Tasks | 65.8 |
| School | 46.0 |
| Hygiene | 44.0 |

**Table 5.** Frequency of Substances Used.

| Substance Used | Frequency of Use (%) |
| --- | --- |
| Tobacco/ Nicotine | 95.2 |
| Marijuana | 92.1 |
| Alcohol | 91.3 |
| Methamphetamine | 90.3 |
| Crack/ Cocaine | 74.3 |
| Opioids | 71.4 |
| Heroin | 67.5 |
| Suboxone | 52.8 |
| Benzodiazepines | 46.0 |
| LSD/ Mushrooms | 45.1 |
| MDMA | 33.1 |
| Inhalants | 18.8 |
| Methadone | 17.2 |
| Other | 15.3 |
| Flakka/ Bath Salts | 12.8 |
| PCP | 10.8 |
| OTC | 7.9 |

## Discussion

The findings from our study identified a notable relationship between participants who screened positive for the Ohio State University Traumatic Brain Injury Identification method (OSU-TBI-ID) and the presence of substance use disorders (SUD). This analysis was conducted using data collected from a substance use disorder treatment program in West Virginia. The lack of adequate training among healthcare professionals in managing these co-occurring diagnoses highlights an urgent need for educational initiatives aimed at enhancing their competencies in treating clients with both TBI and SUD. Future research should prioritize the exploration of integrated treatment approaches for these co-occurring conditions to assess their effectiveness in fostering recovery and improving the overall quality of life for affected individuals.

### Cause of Traumatic Brain Injury

Table 2 displays Motor Vehicle Accidents and Violence as the most reported method of acquiring a TBI among study participants. This is not generalizable on a larger population scale, as the leading cause of TBI in the United States is Falls with Motor Vehicle Accidents as a secondary common cause (CDC, 2024). While falls are related to approximately half of TBI hospitalizations on a national scale, falls were reported as causing only 7.3% of reported TBIs in the study, accounting for 35 out of 483 participants. The disparity in the participant group data and the national data may be attributable to the age range of participants and participants’ history of SUD. Falls on the national level are typically associated with older adults and children, whereas the participant sample for the study primarily comprised individuals within the 18–25-year age bracket. Individuals who experience SUD have a higher association with violence than other populations, which supports the data of Violence being the second largest contributor to TBI among study participants (Schifano et al., 2020).

### Triggers to Use Alcohol and/or Drugs

The National Institute on Drug Abuse (2018) cites work, family, psychiatric illness, pain, or encountering past acquaintances and environments associated with substance use to be the most common potential triggers for individuals experiencing SUD or having a history of SUD. Compared to the results presented in Table 3, family and pain do not have enough significant data points to be displayed in the table for this study. For participants, Work Pressure (22.8%) and Mental Health (62.9%) were not as common as other triggers. Study participants may rank Difficulty Dealing with Feelings (77.2%), Hanging Around Wrong Places (76.0%), and Boredom (70.2%) as the highest occurring triggers for multiple reasons. Individuals who have acquired a TBI are more likely to experience emotional dysregulation, which can cause difficulty in dealing with feelings related to depression, anxiety, and other mental health conditions commonly associated with moderate to severe TBI (Insen et al., 2023). The National Institute on Drug Abuse (2018) identifies environmental cues such as streets, objects, and smells as possible triggers for substance use, which is listed as one of the most common triggers. Hanging Around the Wrong Places, as significantly ranked in Table 3, may include these factors when thinking about what a “wrong” place means to them. The third most frequently seen data point for triggers is Boredom, which may be attributed to the commonly seen loss of employment and inability to participate in once sought-after activities after a TBI has occurred (Alkhawaldeh et al., 2023; CDC, 2024; Khellaf et al., 2019).

### Negative Impact of Substance Use on Daily Functions

Substance abuse can have far-reaching consequences in daily life, particularly affecting personal relationships. According to the results presented in Table 4, a significant number of participants indicated that their relationships were strained because of substance use. This is particularly concerning for individuals living with mental health conditions, as strong relationships are crucial for coping with life’s challenges. Damage to these relationships can hinder feelings of support for individuals with mental health issues, negatively impacting their overall recovery (Chan et al., 2022). As social support diminishes, these individuals may find themselves trapped in a worsening cycle of addiction, seeking solace in substances to fill the void left by lost connections.

Legal issues stemming from drug abuse are also prevalent, with a staggering 89% of participants having faced such consequences. This finding aligns with research indicating that substance abuse frequently leads to legal troubles, including arrests and incarceration (Mackenzie & Styve, 2009; Grossbard et al., 2017). Therefore, integrating education about the legal repercussions of substance use and providing legal assistance within SUD rehabilitation programs are critical steps to mitigate future issues (American Addiction Centers, 2024).

Furthermore, financial difficulties are commonly reported, with 81.6% of participants experiencing economic problems related to their substance use. These financial struggles not only contribute to addiction but are also a significant outcome of drug abuse, highlighting the challenges of achieving sobriety and the impact on overall quality of life. Financial literacy and money management skills should be incorporated into SUD rehabilitation to support individuals in their recovery process (Mattila et al., 2022; Ryan et al., 2023).

Many individuals grappling with mental illness resort to drugs as a coping mechanism. Research shows that substance use leads to a deterioration in well-being for 78.9% of individuals with mental health conditions (McHugo et al., 2017). This statistic underscores the need for holistic treatment approaches that address both mental health issues and substance use. Alarmingly, around 77% of drug abusers report low self-esteem, and 70.6% show poor workplace performance, often resulting in job losses (Awan et al., 2020). Negative self-perceptions are closely linked to increased substance use, reinforcing a cycle of addiction.

Physical health and recreational activities play vital roles in recovery as well, with 69.6% of participants highlighting the importance of physical well-being and 68.7% emphasizing the necessity of leisure activities. Current evidence supports therapeutic systems that incorporate both physical and psychological health, demonstrating a significant relationship between substance abuse and various chronic medical conditions (Miller & Gold, 2019; Haarbauer-Krupa et al., 2021). A lack of meaningful recreational activities can signal reduced quality of life; active engagement in such pursuits is essential for rehabilitation and social reintegration. Additionally, concerning trends are evident, with 65.8% of participants reporting difficulties in daily functioning and 44% neglecting personal hygiene. Substance dependence often correlates with a disregard for self-care and an absence of self-awareness (Centers for Disease Control and Prevention, 2024; Chan et al., 2022). This public health issue can lead to social isolation and increased neglect of existing health conditions.

### Frequency of Substances Used

A comprehensive overview of the prevalence of substance use among individuals with mental health and substance use disorders is provided in Table 5. At the forefront, tobacco or nicotine emerges as the most frequently used substance, with an alarming 95.2% of participants engaging in its use. This finding is consistent with existing research indicating that daily nicotine consumption is prevalent among individuals struggling with mental health issues (Hasin et al., 2013). The social acceptability of tobacco may contribute to its popularity as a stress reliever among this demographic. Following closely, 92.1% of participants reported using marijuana, a trend that can be attributed to its increasing legalization and social acceptance in various regions. Although some individuals turn to marijuana for coping, it is important to note that it can exacerbate certain mental health conditions (Chan et al., 2022).

Alcohol use is another significant concern, with 91.3% of participants acknowledging its consumption, highlighting the pervasive issue of alcohol abuse among those facing mental health challenges (Grossbard et al., 2017). The statistics also reveal a striking 90.3% prevalence of methamphetamine use and 74.3% for crack/cocaine, underscoring the alarming rates of addiction to these substances. The primary drivers behind such drug abuse are often linked to stress or trauma-related experiences (Rosenthal et al., 2023).

Moreover, opioid abuse presents a national prevalence rate of 71.4%, often co-occurring with various mental health disorders. Other substances, such as methadone (46.0%) and benzodiazepines (52.8%), also exhibit considerable levels of abuse among participants. While these medications may initially serve therapeutic purposes, they frequently lead to dependency issues (Khellaf et al., 2019). Additionally, psychedelics like MDMA (18.8%) and LSD/mushrooms (33.1%) are increasingly used recreationally, though their therapeutic effects on mental health remain largely unestablished (Starosta et al., 2021). This complex interplay between substance use and mental health necessitates further exploration and integrated treatment approaches to address these intertwined challenges effectively.

## Limitations and Future Studies

There are some limitations within this study; we are not a part of the initial data collection process with Recovery Point, and the data are taken from multiple different sites. Each site may have a slightly different process, and the assessments that were provided have varying levels of detail. There were potential limitations of human error regarding the data entry process into SPSS as well. There were 96 data points collected per participant, and with 483 participants, the data entry process was completed over 3 months. A limitation was the severity scoring, which indicated withdrawal factors and other symptoms of SUD, but did not explain how they were scored when going through the intake. A limitation in categorizing drug usage is that the duration or frequency of drug usage was not indicated in this research, which could skew certain drugs that were only used once or are substances they no longer use. Another limitation was in categorizing education in this research because it was only classified by degrees and not some college experience, vocational programs, or certifications. This could result in an inaccurate representation of the education level of the participant because it will only show that they have a high school diploma and no other experience.

Future studies should include more qualitative data collection to expand further upon the triggers and the negative impacts on daily function. Having their thoughts instead of a curated list of options would provide a better understanding of the individual. Emphasizing how their TBI or undiagnosed TBI has impacted their daily function to better understand this relationship. Future studies could analyze the negative screeners and compare them to the positive ones to see if TBI has more of an impact on these factors. Future studies could see if there is an increased correlation between the positive group for substances, triggers, and negative impact because it could be seen across all SUD diagnoses.

## Conclusion

### Circle Figure 1

This study aimed to investigate the interaction between traumatic brain injury (TBI) and substance use disorder (SUD) in participants in a substance use program. We found that TBI is quite common in people with SUD and that having a TBI seems to predispose people to engage in risky behaviors, which then puts them at higher risk for substance use. On the other hand, SUD can worsen the cognitive and emotional difficulties that come with TBI.

To re-examine the research question, the study aimed to determine whether SUD leads to TBI or vice versa. What was discovered was that the two conditions are indeed interconnected; people who have TBI can use substances as a coping mechanism, therefore indulging in risky activities and becoming susceptible to obtaining TBI. What’s interesting about what we discovered is that it highlights the importance of treating TBI and SUD simultaneously in medical settings. Understanding how these two conditions relate to each other is the key to creating effective treatment plans that address both conditions simultaneously.

By treating both together, medical professionals can help those with these dual issues thrive, eventually leading to a better quality of life and increased independence. With that said, we do have to acknowledge that there are some limitations to our study. For example, relying on self-reported information can be somewhat prejudiced, and data in our study were taken at a snapshot in time, so we can’t always assume cause and effect.

Future studies should utilize long-term research and broader, more diverse study groups to assess how TBI and SUD intersect in the long term. We encourage more focused research to develop targeted therapies specifically for individuals experiencing both SUD and TBI. As we learn more about the conditions, we suggest clinicians embrace evidence-based practices that advance quality of life, enable long-term rehabilitation, and meet today’s urgent problems. Ultimately, the complex dynamic between TBI and substance use disorder offers fertile ground for improved outcomes. With repeated practice and enhanced education for clinicians, we can treat this risk population of patients more effectively with successful rehabilitation and recovery.

## Data Availability

All data produced in the present study are available upon reasonable request to the author Lura Leech

